# A systematic review of Crimean-Congo Haemorrhagic fever virus in Sub-Sahara Africa, 1969-2022

**DOI:** 10.1101/2022.10.28.22281642

**Authors:** Joseph Ojonugwa Shaibu, Olufemi B. Daodu, Kabiru Olusegun Akinyemi, Rosemary Ajuma Audu, Akeeb O. Bola Oyefolu

**Affiliations:** Department of Microbiology, Faculty of Sciences, Lagos state University, Ojo, Lagos, Nigeria; Centre for Human Virology and Genomics, Nigerian Institute of Medical Research, Yaba, Lagos State, Nigeria; Department of Veternary Medicine, University of Ilorin, Kwara State

**Keywords:** Haemorrhagic, PRISMA, seroprevalence, ELISA, Sub-Saharan, Clade

## Abstract

In Sub-Saharan Africa, CCHFV has been reported in some of the countries with resultant seroprevalences ranging from 1.65-44%, 0.37-75%, 19-74% amongst humans, ticks and cattle respectively using ELISA technique. Strains classified as Africa clade 1,2 and 3 have been established to be circulating in Sub-Saharan Africa from few molecular studies carried out. CCHFV has part of its nomenclature rooted in Africa, however, the actual spread of the virus across sub-Saharan Africa is poorly known. This paucity of knowledge is attributed to less work done in the quest to understand this virus better. Most researches, which were quite limited, carried out across Africa were on antibody detection using ELISA methods with little information on molecular characterization. The aim of this review is to harmonize the studies carried out in Sub-Saharan Africa on CCHFV between 1969 -2022 with respect to seroprevalence, viral identification and isolation, molecular characterization and genomic analysis. Articles are retrieved from public databases such as google search, PubMed, Google scholar and ResearchGate, filtered using PRISMA concept and data extracted from eligible articles and analyzed. In this study the overall average seroprevalence of CCHFV in Humans, Ticks, Cattle, sheep and Goats are 13.6%, 18.1%, 44.3%, 11.3%, 16.3% respectively. South Africa has the highest seroprevalence (20.8%) among humans and Uganda (2.5%) has the lowest. The prevalence of CCHFV in many African countries is still yet unknown though there is clear evidence of exposure of people within the region to CCHFV. Limitations in sensitivity and specificity of diagnostic techniques such as agar gel precipitation test, haemagglutination test and complement fixation test used at some instance suggest a need for more reliable techniques.

**Author’s summary:** Crimea-Congo haemorrhagic fever virus is carried by ticks. It has a high fatality rate among humans. It is implicated in haemorrhagic fever with bleeding through the nose and mouth. CCHFV is fast spreading across the world but little is known it in Sub-Saharan Africa. Many individuals in this region are herders, they are daily exposed to ticks; they regularly come down with febrile illnesses that are most times misdiagnosed as either malaria or typhoid. In the course of treating for malaria or typhoid, some die. There is no awareness ongoing in the communities about this infection and its danger to the population, preventive measures with respect to personal hygiene, cleanliness of the abattoirs and general environments and the need to seek medical attention and avoid self-medication. In this review it is shown that there is evidence of spread of CCHFV in Sub-Saharan Africa, however, there is paucity of information. This lack is as result of many factors such as lack of funds for research, porous security that makes it difficult tracing, lack of effective diagnosis of viral pathogens and so on.

## INTRODUCTION

Crimean-Congo haemorrhagic fever virus (CCHFV) is among the highly priority pathogens considered by World Health Organization research and development Blueprint (WHO R& D) based on its high case fatality rate, tendency for hospital outbreaks and challenges involved in treatment, control and prevention **[1]**. It belongs to: **Realm:** Riboviria; **Kingdom:** Orthornavirae; **Phylum:** Negarnaviricota; **Class:** Ellioviricetes; **Order:** Bunyavirales; **Family:** Nairoviridae; **Genus:** Orthonairovirus; **Species:** Crimean-Congo hemorrhagic fever orthonairovirus. CCHFV is widely spread and presently reported in Africa (Nigeria, Kenya, Ghana, Mauritania, Cameroun, South Africa), Asia (Xin Jiang region of China, the Middle East and Southern Russia), Europe (southern and eastern parts), the Middle East and the Indian sub-continental **[2-4]**. Globally, more countries of the world are reporting its incidence and there is currently no internationally licensed vaccine for human or animal use. Crimean-Congo haemorrhagic fever virus (CCHFV) is implicated in severe haemorrhagic disease in humans but transient pyrexia in infected livestock (cattle, sheep and goats) [1]. After Dengue fever, CCHFV is reported to be the second widely distributed agents of severe haemorrhagic fever known [5].

It is classified along with other viruses in the families *Arenaviridae, Filoviridae, Bunyaviridae, Flaviviridae*, and *Rhabdoviridae* as viral haemorrhagic fever based on common signs of fever and haemorrhage in the infected animal and or humans.

It causes up to 50% mortality in humans **[1]**. CCHF is a notifiable disease to the World Organization for Animal Health due to its ease of spread and clinical implications in humans. In Sub-Saharan Africa, a lot is yet to be known about CCHFV and its implication on public health. Just like many other arboviruses, there is a lot of misdiagnoses of every febrile illness for malaria or typhoid and this is partly due to lack of resources and technical ability to conduct researches that can enlighten the public about possible pathogens circulating the society and their signs and symptoms. This in turn helps policy making by the government to effectively manage disease outbreak. Checking through public databases, there is a paucity of knowledge on CCHFV and other VHF viruses of public health importance in Africa. Hence, the aim of the review is to harmonize studies that have been conducted in Sub-Saharan Africa in order to give at a glimpse, idea on gaps that need to be covered.

## METHODOLOGY

Original articles and review papers carried out in Sub-Saharan Africa in public databases such as NCBI, PubMed, Google scholar, Google search, Research gate were searched using statements such as “Crimean-Congo Haemorrhagic Fever Virus in Sub-Saharan Africa” OR “Seroprevalence of CCHFV in Sub-Saharan Africa (Nigeria, Ghana, Gambia, Cameroun, Senegal, Niger, South Africa, Malawi, Mauritania, Mozambique, Zambia, Sudan, Tanzania, Republic of Guinea, Cote d’Ivoire, Uganda etc.” OR “Characterization of CCHFV in Sub-Saharan Africa” OR “History of CCHFV” were retrieved and reviewed to extract information on the seroprevalence, molecular characterization, sample size, source of sample, location of study, risk factors, diagnostic method, etc.

### INCLUSION CRITERIA

Laboratory studies that reported prevalence and/or characterization of CCHFV in Sub-Saharan Africa using serological, cell isolation and/or molecular techniques were included in this review. Research focusing on Humans, Ticks, cattle, Goats, sheep and other animals were included.

### EXCLUSION CRTERIA

This review excludes any article on CCHFV which was not lab-based. It excludes reports on other viral haemorrhagic fever viruses. Studies conducted on CCHFV outside Sub-Saharan Africa were excluded as primary focus of the review.

### DATA EXTRACTION

Based on the objective of this study, relevant quantitative and qualitative data were extracted from journals that met the eligibility criteria. Information on parameters such as; study location, period of study, sample size, sample source, type of sample, diagnostic method, risk factors, prevalence, case fatality, strain and treatment method were extracted using Microsoft excel 2019 workbook with each column representing each of the parameters. Data were cleaned and then analyzed.

### RESULTS

The online search on the public databases yielded 1610 articles which had the name of the pathogen – CCHFV. Further screening of the journals to match the eligibility criteria for this review eventually yielded 32 articles (Fig 1) which were used for this review. Results were extracted from 18 countries which conducted studies that met the criteria (Table 1).

**Fig 1:**
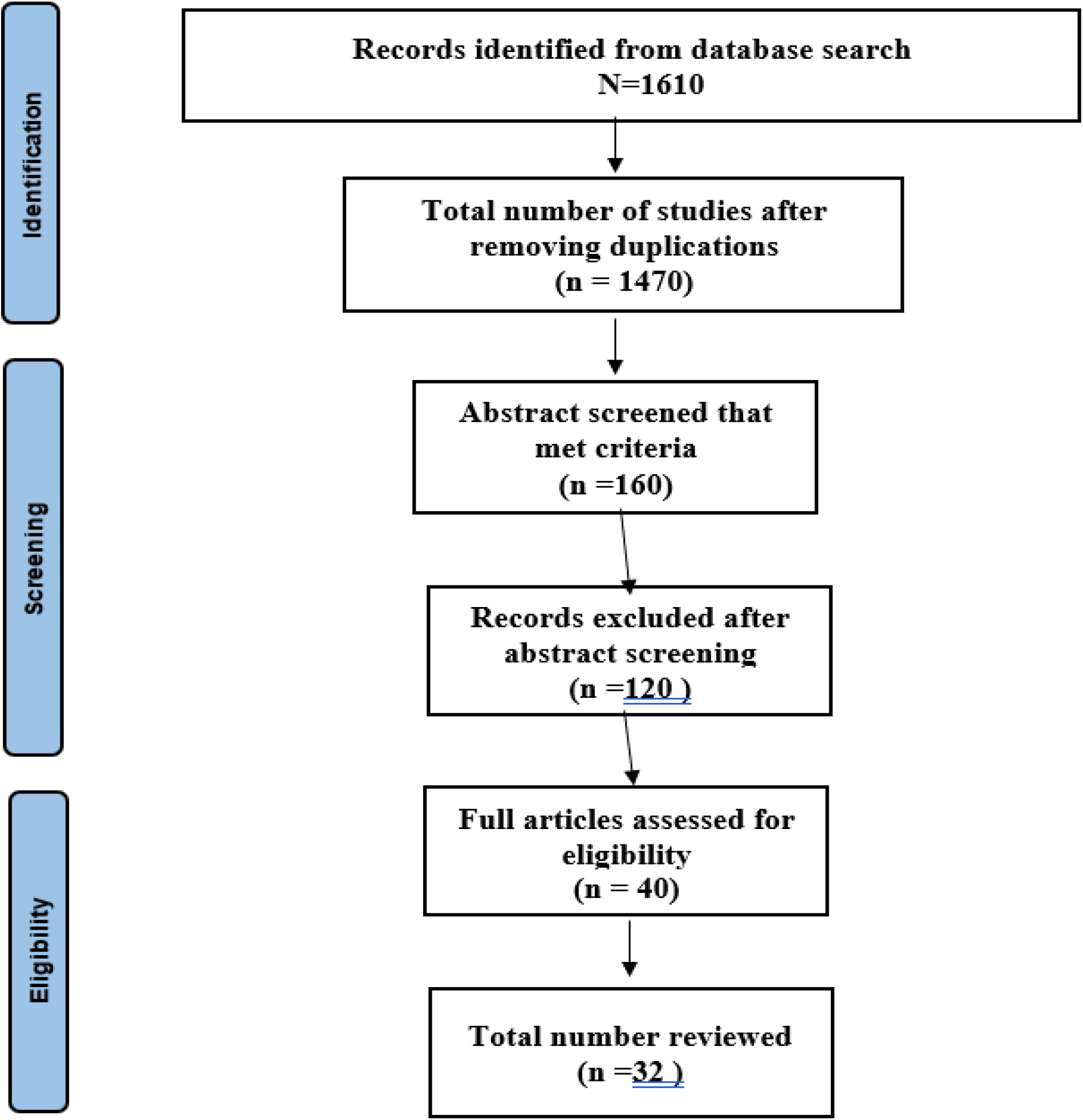
Adapted PRISMA flow chat.

**Table 1:**
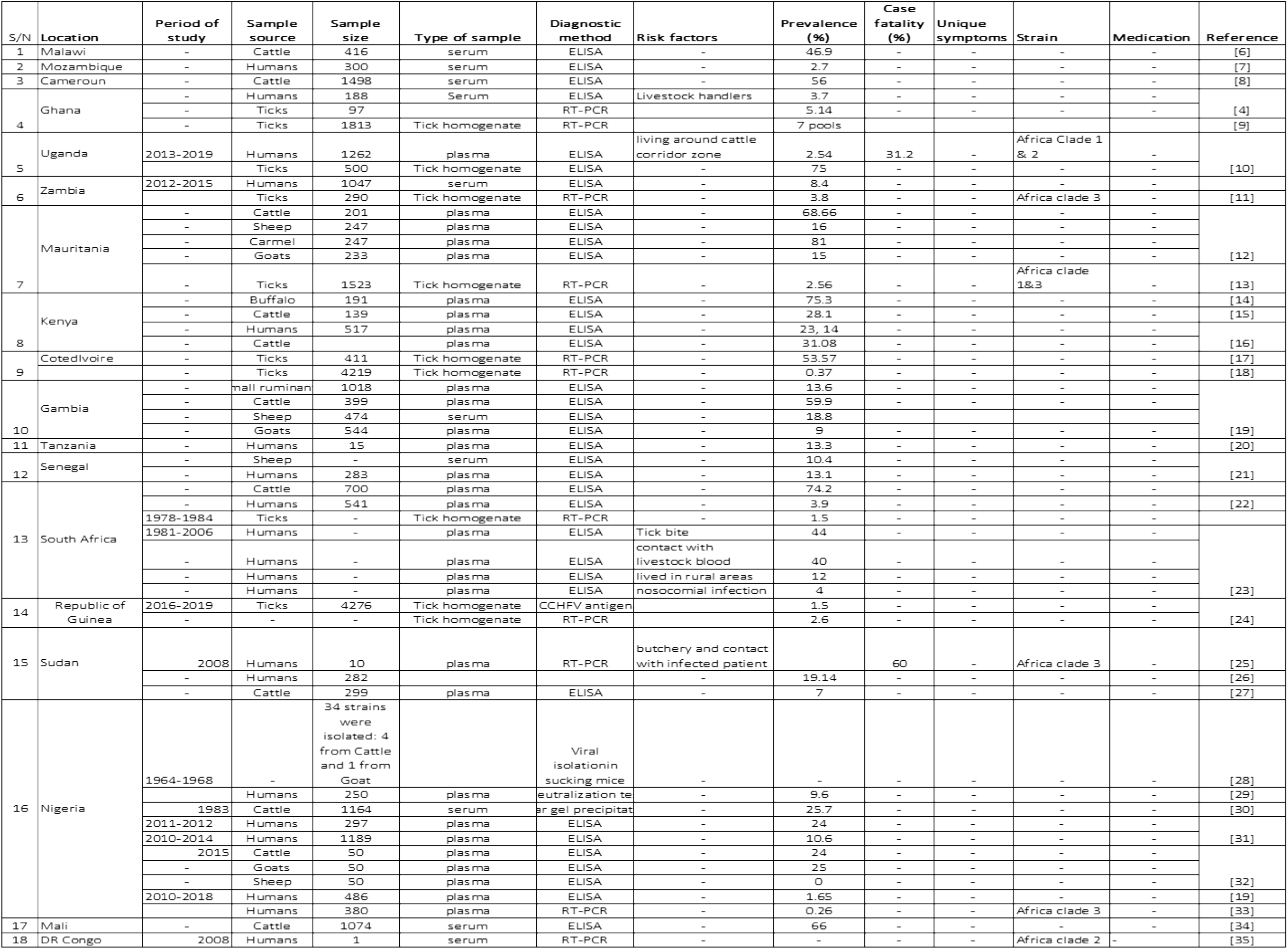
Summary of original studies conducted on CCHFV across Sub-Saharan Africa.

## SEROPREVALENCE OF CCHFV ACROSS SUB-SAHARAN AFRICA

In this study the seroprevalence of CCHFV as researched in different countries ranges between 0.26-75% (Table 1). However, the overall average seroprevalence of CCHFV in Humans, Ticks, Cattle, sheep and Goats are 13.6%, 18.1%, 44.3%, 11.3%, 16.3% respectively (Table 2) across eighteen (18) countries. Across Sub-Saharan Africa, the study subjects were 40% on humans, 18% on Ticks, 22% on Cattle, 8% on Sheep, 6% on Goats and 6% on other animals such as Carmel and Buffalos (Fig 2). The percentage usage of ELISA technique in the serostudies reviewed was 63.2% while the use of RT-PCR, sequencing and other serological methods were 19.3%,10.5% and 7% respectively (Fig 3).

**Table 2:** Seroprevalence of CCHFV among selected sources across Sub-Saharan Africa.

| **Study subject** | **Seroprevalence (%)** |
| --- | --- |
| Humans | 13.6 |
| Ticks | 18.1 |
| Cattle | 44.3 |
| Sheep | 11.3 |
| Goats | 16.3 |

**Fig 2:**
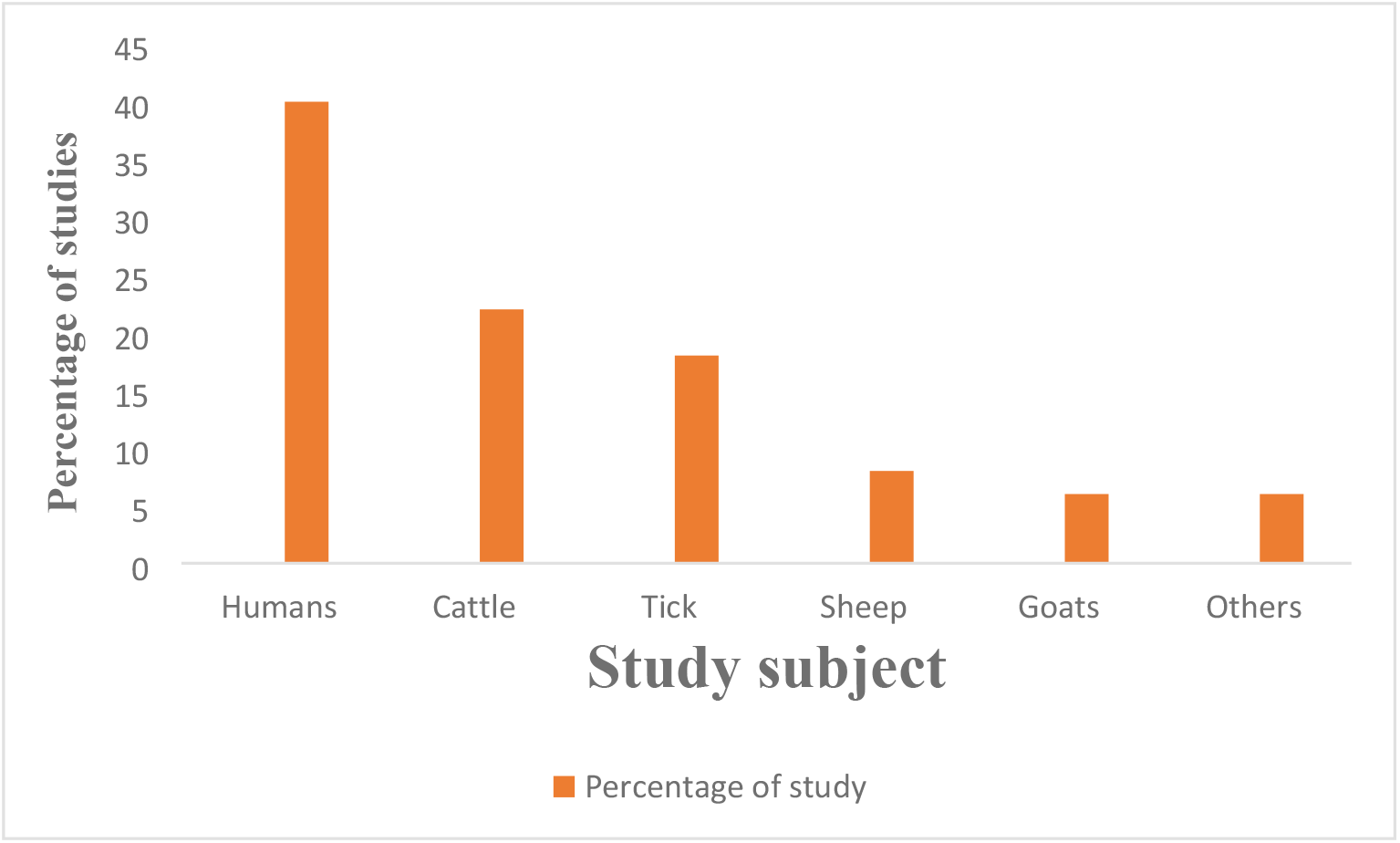
Percentage study per study subject.

**Fig 3:**
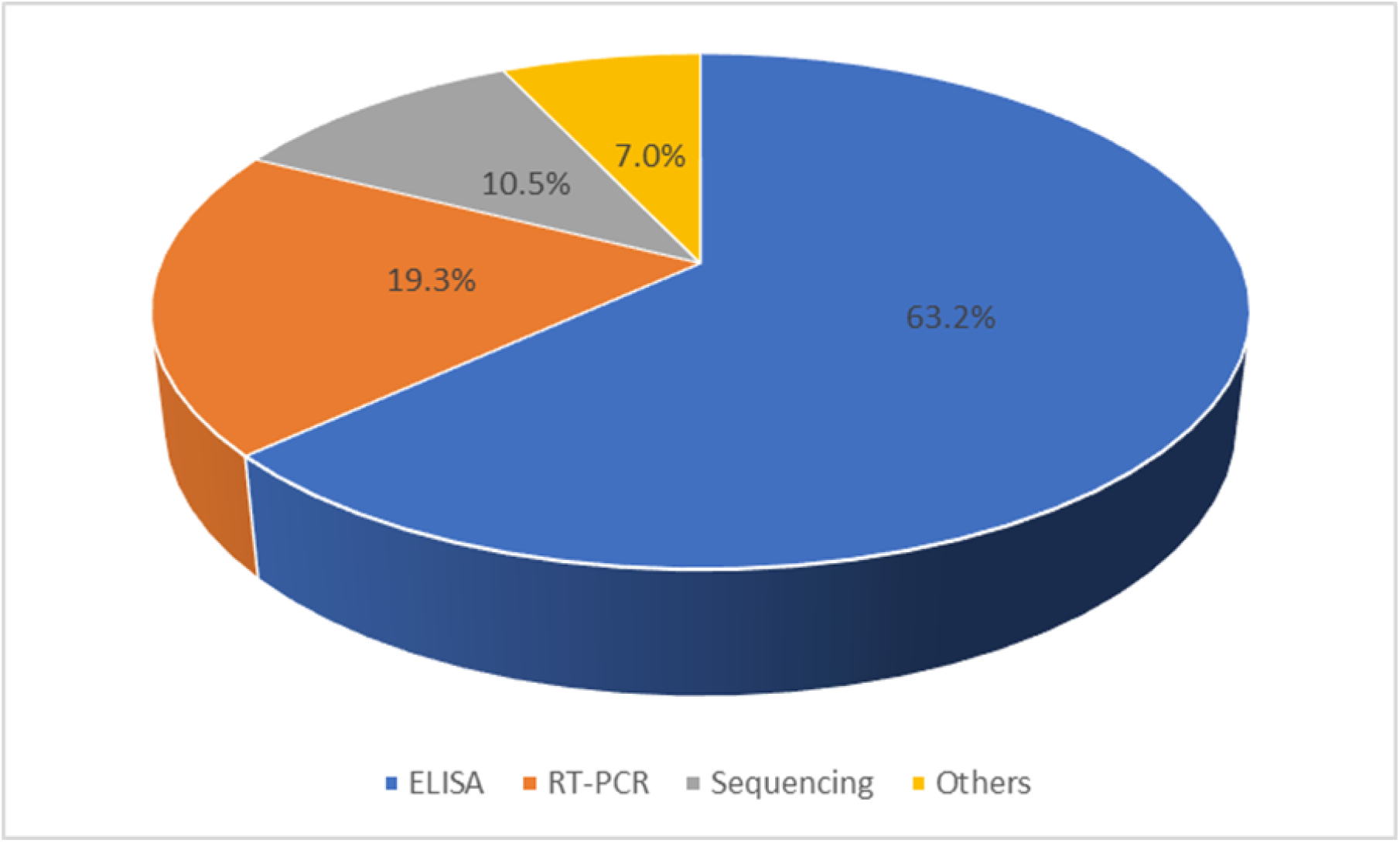
Pie chart on the study techniques used for CCHFV study across Sub-Saharan Africa.

Comparing seroprevalences across the countries in this study, it can be seen from Table 3 that South Africa has the highest seroprevalence (20.8%) among humans and Uganda (2.5%) has the lowest. Nigeria has an average seroprevalence of 11.5%, Kenya 18.5% and Sudan 19.1%.

**Table 3:** Summary of seroprevalences of CCHFV across some countries in Sub-Saharan Africa.

| S/N | Country | Seroprevalence |  |  |  |  |
| --- | --- | --- | --- | --- | --- | --- |
|  |  | Human (%) | Tick (%) | Cattle (%) | Sheep (%) | Goat (%) |
| 1 | Malawi | - | - | 46.9 | - | - |
| 2 | Mozambique | 2.7 | - | - | - | - |
| 3 | Cameroun | - | - | 56 | - | - |
| 4 | Ghana | 3.7 | 5.1 | - | - | - |
| 5 | Uganda | 2.5 | 75 | - | - | - |
| 6 | Zambia | 8.4 | 3.8 | - | - | - |
| 7 | Mauritania |  | 2.6 | 68.7 | 16 | 15 |
| 8 | Kenya | 18.5 | - | 29.6 | - | - |
| 9 | Cote D Ivoire | - | 53.6 | - | - | - |
| 10 | Gambia | - | - | 59.9 | 18.8 | 9 |
| 11 | Tanzania | 13.3 | - | - | - | - |
| 12 | Senegal | 13.1 | - | - | 10.4 | - |
| 13 | South Africa | 20.8 | 1.5 | 74.2 | - | - |
| 14 | Republic of Guinea | - | 2.6 | - | - | - |
| 15 | Sudan | 19.1 | - | - | - | - |
| 16 | Nigeria | 11.5 | - | 24.9 | - | 25 |
| 17 | Mali | - | - | 66 | - | - |

## MOLECULAR CHARACTERIZATION

Out of eleven studies carried out using RT-PCR for the prevalence of CCHFV, only 6(54.5%) proceeded to sequencing. Of the 6 samples analyzed, 1(16.7%), 1(16.7%),3(50%), 1(16.7%) were grouped under Africa clade 2, Africa clade 1 & 2, Africa clade 3 and Africa clade 1 & 3 respectively.

Out of 48 Sub-Saharan African countries, by the criteria of this review, only 18 (37.5%) countries have carried out a study using either serology and/or molecular technique (Fig 4).

**Fig 4:**
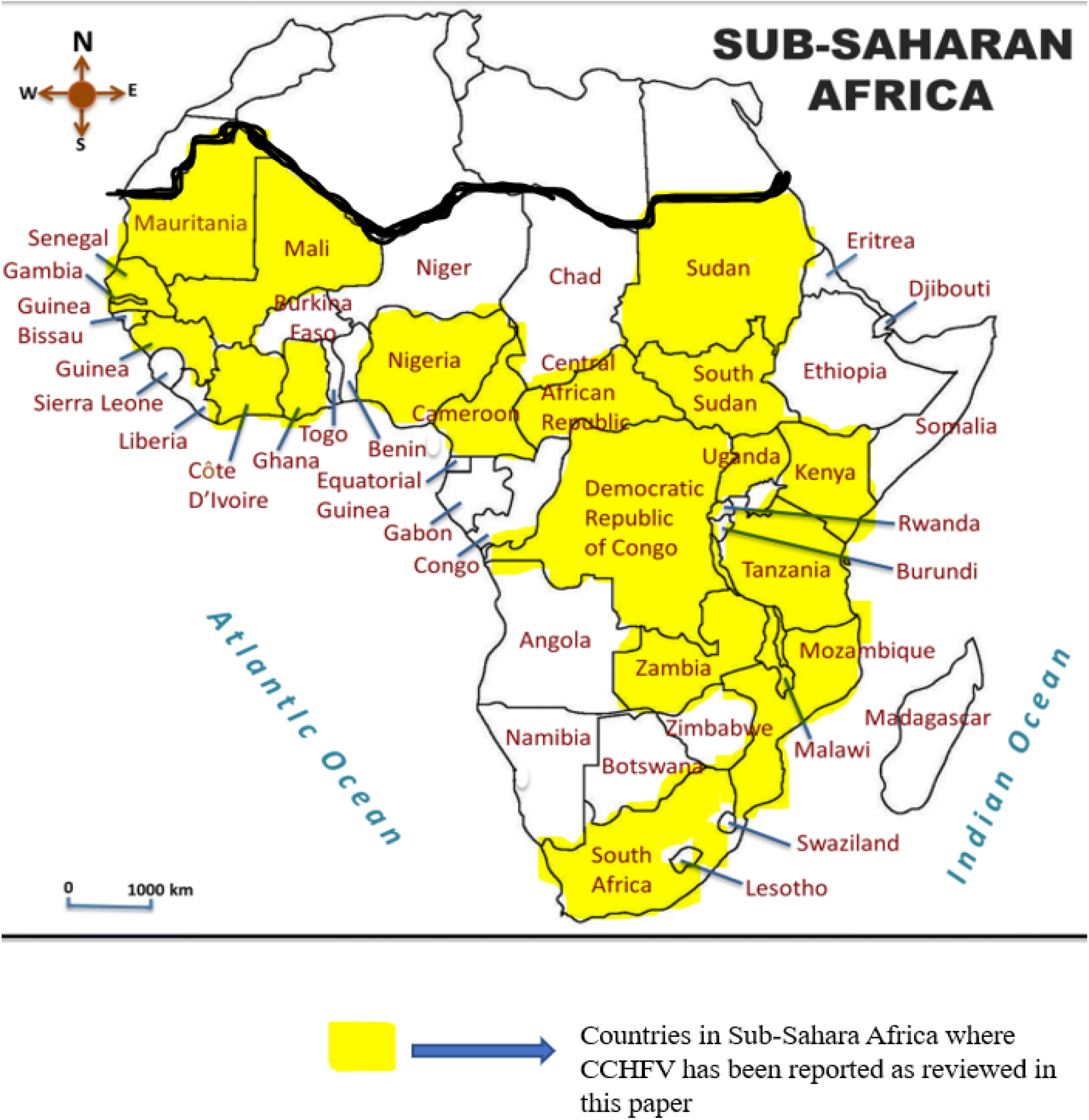
Map of Africa showing distribution of CCHFV in Sub-Saharan Africa.

## DISCUSSION

In the past few years, zoonotic infections are on the increase due to the increasing interactions between man, animals and the environment. This has given room to the rising of many emerging and re-emerging pathogens. CCHFV is an emerging pathogen capable of causing high fatality in humans.

Studies on CCHFV in Sub-Saharan Africa is highly limited considering the number of countries in this region and the fact that the virus partly had its history here. This paucity of knowledge is majorly as a result of lack of or low-level research work been done on CCHFV in Sub-Sharan Africa. This is contrary to many countries in Asia and Europe where a lot of work has been done providing a better insight about the virus in those regions [36]. Possible identifiable causes of this limitation in research on CCHFV could arise from:

1. Lack of funding for research into pathogens of public health importance: this fate applies to majority if not all of the countries in Africa; there is no funding allocation from governments towards research and even from private organizations. Hence, researchers in this region heavily depend on collaborations and grants from international organizations which are not easy to come by. This rigour of getting funds for research makes many scientists in Africa become redundant. To help solve this problem, African government need to make funds available for research works into diseases of public health importance in order to reduce mortality from infectious pathogens.
2. Cost and availability of diagnostic tools: Diagnostics tools (both in equipment and reagents) are far-reached by scientists in this region, hence the passion for consistent, effective and productive research works is lost. The over-dependence on foreign kits for diagnosis in unstable economies as seen in Africa makes cost of purchase so high because of constant increasing difference in currency exchange rates. Without sensitive and specific diagnostic tools, research is cumbersome as results obtained otherwise are not reliable. Most illnesses especially resulting in fever are quickly empirically diagnosed as either Malaria or Typhoid fever by mere observation and no laboratory confirmation [37,38,39]. It is advisable for private institutions to rise in collaborations with scientists to develop in-country diagnostic kits that will also be suitable for the diagnosis of the various strains of pathogens circulating in Sub-Saharan Africa.
3. Security: Most “VHF” researches involve collection of clinical samples from field work; several times, this is faced with lots of challenges as a result of poor security situations in many African countries. For instance, in a study reported by Aradaib *et al* on an outbreak of CCHFV in Sudan, there was no effective contract tracing to track all the people possibly infected because of security challenges. Also, in Nigeria, a study by [33] reported the first human case of CCHFV in Nigeria but did not give any information on contact tracing, this is assumably as a result of security challenges in that region.

From this study, the average seroprevalence of 13.56% recorded in humans showed that there is possibly a high level of exposure of people to CCHFV in Sub-Saharan Africa. However, this rate when compared to the seroprevalences (18.1%, 44.3%, 11.3%, 16.3%) of the same pathogens in the various hosts (Ticks, Cattle, Sheep, Goats) respectively (Table 2) confirms that the study coverage among humans is low since there is constant interaction between these animals and humans. This is because a large population of individuals in Sub-Saharan Africa are herders and butchers who daily interact with these group of animals, hence there should be higher level of exposure to infected ticks and the animal hosts. Also, there is constant movement of flocks and their herders across several African countries; the herders interacting with people in publics places such as markets and religious gatherings thereby exposing more persons to possible infection of CCHFV. On individual country bases, South Africa with an average seroprevalence of 20.8% seems to be the highest across Sub-Saharan Africa. This could be interpreted with caution since they have carried out more serostudies for CCHFV among humans compared to other countries and they may have better and applicable diagnostic facilities thereby generating more data than other countries in the region. Evidence of the exposure of other animals such as Carmel, Buffalos and bats [12,14] signifies that there could be many more hosts yet unknown. There is need for more serological studies to provide robust information on the true burden of CCHFV among humans in Sub-Saharan Africa.

Looking at the methods of diagnosis used in the various studies analyzed in Table 1, 63.2% were IgG screening based. This method only detects previous exposure in which the pathogen might not be actively present in the sample at the point of analysis. Hence, for detection and characterization of the virus, molecular studies are necessary [40]. This provides more information on the strain of the pathogen in circulation, virulence and pathogenesis of the circulating strain and possible cluster of cases within the community. Unfortunately, from the journals reviewed, only 19.3% of the studies were performed using RT-PCR, however, among these, only 54.5% proceeded to sequencing level hence identification of the strains involved. This dearth in molecular study within the region drastically reduces the chances of development of effective diagnostics, drugs and vaccines that will be useful for the management of diseases arising from CCHFV infections specific to the strains circulating in the region. Lack of sequence data on CCHFV in public databases from Sub-Saharan Africa limits the consideration of the strains in circulation in the region in the development of diagnostics and therapeutics, hence the region will be limited to products developed using strains circulating in other regions of the world which sometimes give detection failures in Sub-Saharan Africa. This is arguably one of the reasons why the etiology of most fever cases is still unknown in Sub-Saharan Africa [38].

The case fatality of 31.2 as reported by [10] from a study conducted in Uganda among humans is a major public health concern. Unfortunately, because of limited studies on humans and non-effective case tracking in the studies reported, more data are needed to define case fatality of CCHFV across Sub-Saharan Africa.

## CONLUSION

From the review, it is clear that there is exposure to CCHFV in Sub-Saharan Africa, much is not known yet about the virus and how it affects man in Sub-Saharan Africa. Hence, there is a need for a wholistic research approach to understand the characteristics of the virus in this region.

## Data Availability

All data have been included

## ACKNOWLEDGEMENT

I want to appreciate all members of staff of the Department of Microbiology, Lagos State University (LASU), Ojo, Lagos State, Nigeria for their contributions to this manuscript.

